# A Comparison of Covid-19 Patient Characteristics Before versus After Partial Lockdown in Vietnam

**DOI:** 10.1101/2020.08.27.20183616

**Authors:** Hoang-Long Vo, Kim-Duy Vu

## Abstract

**Background:** Given the Covid-19’ sudden rapid spread in the community since 25 July 2020, an updated analysis of Covid-19 cases was conducted to examine the differences of characteristics of Covid-19 patients before versus after partial lockdown end in Vietnam.

**Methods:** Data of 569 Covid-19 patients confirmed SARS-CoV-2 infection from 23 January to 31 July was collected from available official databases. We divided Covid-19 situation timeline into two main periods, before lockdown end (23 January – 22 April) and after lockdown end (23 April – 31 July).

**Results:** We found significant variations in the distribution of Covid-19 patients among different provinces between two periods. Covid-19 confirmed patients were older in the time after lockdown end compared to in the period before lockdown end by a median of 5 years. All discharged Covid-19 patients and no Covid-19 death were in the phase before lockdown, while post lockdown period had still remained significant patients being under treatment, especially reported first fatalities. The number of Covid-19 patients who returned from other countries (excluding China) slightly increased through two stages (p >0.05), partially showing that the continuous volume of people returning Vietnam from abroad during the Covid-19 epidemic.

**Conclusions:** Our analysis indicated demographic and epidemiological disparity of Covid-19 patients before versus after loosening the national partial lockdown in Vietnam. It is important to suggest that, proactive efforts in Covid-19 control after partial lockdown end will be effective when the measures to closely control and monitor repatriation and immigration via the borders of Vietnam are strictly enforced.

**What is already known on this topic:**

- Most of Covid-19 confirmed patients in Vietnam were acquired overseas.
- In Vietnam, Covid-19 epidemic had the low estimated reproduction ratio (R_0_) and SARS-CoV-2 spread was on the downward trend before July 25.

**What this study adds:**

- Covid-19 confirmed patients were older in the time after lockdown end compared to in the period before lockdown end by a median of 5 years.
- All discharged Covid-19 patients and no Covid-19 death were in the phase before lockdown, while post lockdown period had still remained significant patients being under treatment, especially reported first fatalities.
- Continuous volume of people returning Vietnam from abroad through two stages during the Covid-19 epidemic.
- Our analysis might be considered as a prompt reference source for further in-depth surveys to understand the adaptive models of Covid-19 patients among different provinces.

## Introduction

Since the beginning of the coronavirus disease 2019 (Covid-19) crisis, the lockdown has been considered a key policy response in many countries to prevent the further spread of the severe acute respiratory syndrome coronavirus 2 (SARS-CoV-2) which causes the disease. The implementation of the lockdown order, however, has varied in countries and territories around the world, thanks to the unpredictable situation of the pandemic [1]. Vietnam is one of the few countries achieving effective control of Covid-19 outbreak after the implementation of lockdown strategy [2]. Under Directive No. 16/CT-TTg issued in March 31, 2020, a “stay-at-home” order was imposed nationwide to curb the spread of the contagion, limit the number of infections and casualties, and alleviate the pressure on the health service providers. The effectiveness of such outbreak control was evidenced by the fact that there were only 59 new infected cases confirmed during the first 14-day nationwide social distancing (from April 1 to 14, 2020), equaled to 40% of the two weeks prior to the national lockdown [3]. Of these, there were 30 confirmed cases in isolation zones and 29 confirmed cases in the community [3]. Within 99 days since April 15, Vietnam had confirmed no community transmission despite extensive testing. While in many regions of the world strict Covid-19 lockdown orders had still been in effect, the Government of Vietnam relaxed social distancing rules for almost all provinces and cities on 23 April [4]. The early success in Vietnam’ epidemic control has mainly attributed to the national emergency response across the whole sociopolitical system, in particular, 22-day nationwide lockdown.

However, the strike of no Covid-19 community transmission has been broken on July 25 when a new case were reported in Da Nang City [5]. This disruption made the Covid-19 pandemic in Vietnam more unforeseeable than ever [6], [7], [8]. Immediately, contact tracing efforts as well as epidemiologic investigations have been strengthened under the strong guidance of the Vietnamese Prime Minister, yet the source of the infection is not known in several new cases [8], [9]. With the estimation of the reproduction ratio (R_0_) for 257 first Covid-19 patients in Vietnam (between 23/1/2020 and 10/4/2020), a Huy G. Nguyen’ early analysis revealed the downward trend of the epidemic in this country [10]. However, the potential SARS-CoV-2 transmission in the community is not reflective of current reality in each stage under Government’ strict restrictions. An initial understanding of the characteristics of Covid-19 patients detected before and after lockdown end in Vietnam is needed to suggest prompt and efficient actions in the worst-case scenarios for in Vietnam. Given the Covid-19’ rapid spread, an updated analysis of cases was conducted to examine the differences of characteristics of Covid-19 patients before versus after lockdown end in Vietnam.

## Methods

In this paper, we collected data of Covid-19 patients who were confirmed SARS-CoV-2 infection from 23 January to 31 July, 2020. We divided Covid-19 situation timeline from 23 January to 31 July 2020 into two main periods, before lockdown end (23 January – 22 April, 2020) and after lockdown end (23 April – 31 July, 2020). We selected cut-off point of Covid-19 situation timeline in Vietnam on 23 April, 2020 (23 April, 2020 is the day that the Vietnam Government decided to loosen the national lockdown [4]).

### Data sources

Analysis data in this paper was extracted from two sources, official database of the Ministry of Health of Vietnam (MOH) (https://ncov.moh.gov.vn/) and the website of Our World in Data (https://ourworldindata.org/coronavirus#coronavirus-country-profiles) [11]. A total of 569 Covid-19 confirmed patients were consecutively detected from 23 January to 31 July, 2020 (Figure 1).

**Figure 1.**
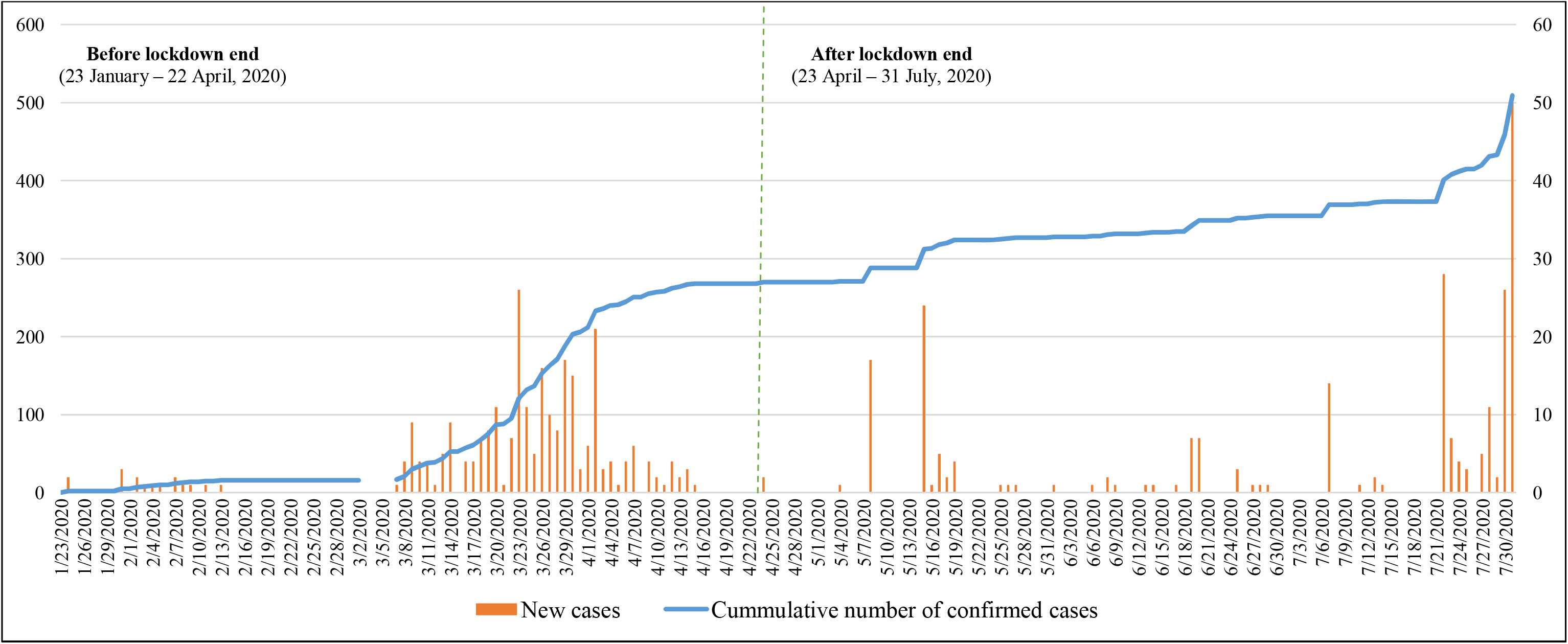
The incidence of confirmed Covid-19 cases and its cumulative numbers, 23 January to 31 July 2020

### Case definition for Covid-19

*Covid-19positive case* and *Covid-19 suspected case* were defined according to the Decision No. 322 / QD-BYT of February 6, 2020 of the Minister of Health for the period before lockdown end [12] and according to the Decision 1344 / QD-BYT 2020 Guidance on diagnosis and treatment of COVID-19 [13] for the period after lockdown. The Vietnam’ tests were reported by the WHO and MOH approved laboratories [12], [13].

*Covid-19 recovered case* before and after lockdown end was defined that, a patient who recovered from Covid-19 and discharged from the health facility met the following criteria: 1) no fever for at least three days; 2) a good general health condition; (3) two samples taken at least 1 day apart having negative test results for SARS-CoV-2 [12], [13].

*Covid-19 death case* is a patient with confirmed Covid-19 infection whose death resulted from clinically compatible illness, unless there is an other certain cause of mortality that cannot be related to Covid-19 disease (e.g., trauma). No period of full recovery was recorded between the illness and death.

### Variables

The variables in this analysis were selected on the basis of the availability of above official databases. The variables related to the Covid-19 patient characteristics were divided into subgroups as following: mean age, age group (0-10 years/11–20 years/21–30 years/31–40 years/41–50 years/51–60 years/60 years and over), gender (male/female), nationality (Vietnam/others), the history of returning from China (yes/no), the history of returning from other countries (excluding China) (yes/no), and treatment status (under treatment/ discharged/ died).

### Data analysis

Vietnam’ Covid-19 situation timeline was considered as a variable with two main values: (code 1) before lockdown end and (code 2) after lockdown end. We used both descriptive and analytical method. While age data was expressed as mean ± standard deviation (SD) with interquartile ranges (IQR), categorical variables were presented as frequency with percentage (%). We estimated the overall proportion of Covid-19 confirmed cases and its percentage before and after lockdown end according to selected demographic and epidemiological characteristics. Chi-square tests and Mann–Whitney U tests were utilized to compare patient’ characteristics between two periods. All statistical analyses in this paper were conducted by Stata® 15 (StataCorp LLC, USA). The level of statistical significance was set at 0.05.

## Results

Among the 569 patients from 23 January to 31 July 2020, there were 268 patients by 22 April and 301 patients between 23 April and 31 July (Figure 1). Figures 2 illustrated the significant variation in the distribution of Covid-19 patient among different provinces between two periods.

**Figure 2a.**
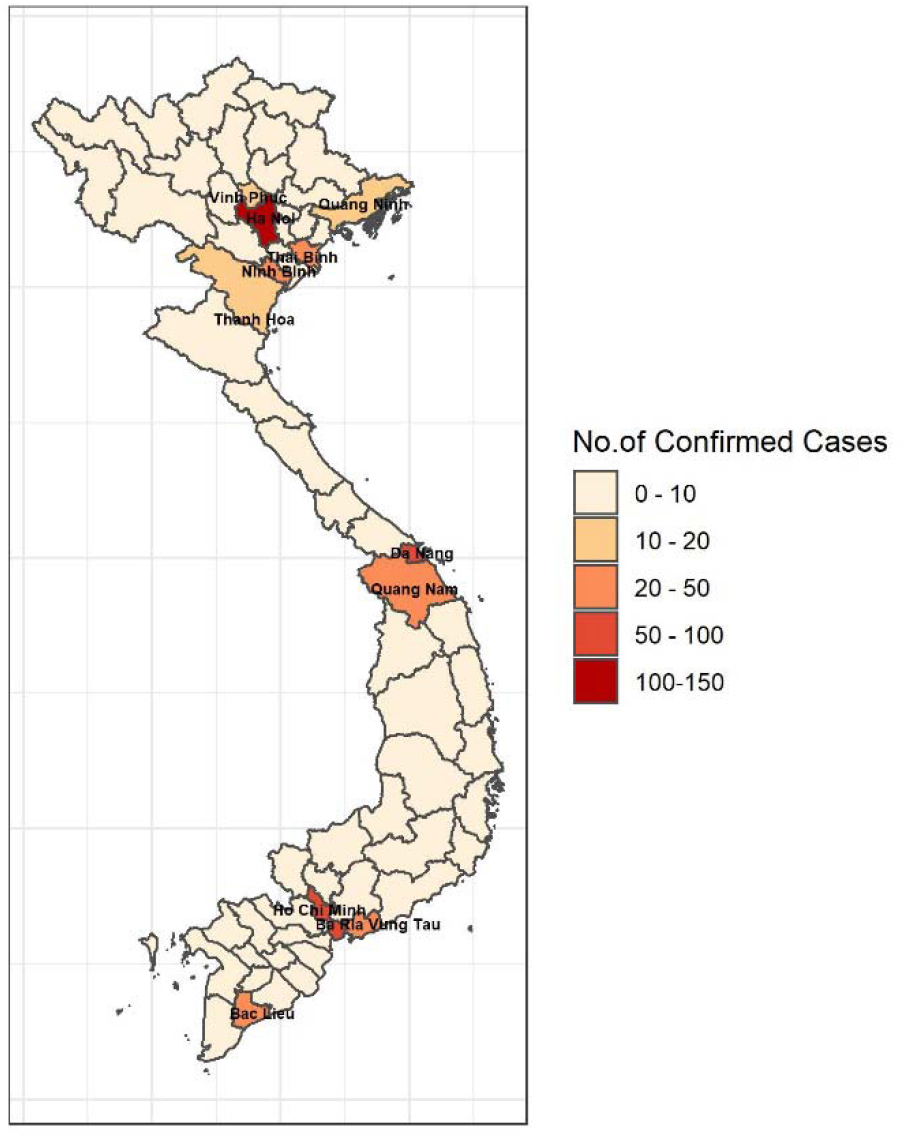
Distribution of Patients with Confirmed Covid-19 Cases across Vietnam by 31 July 2020

**Figure 2b.**
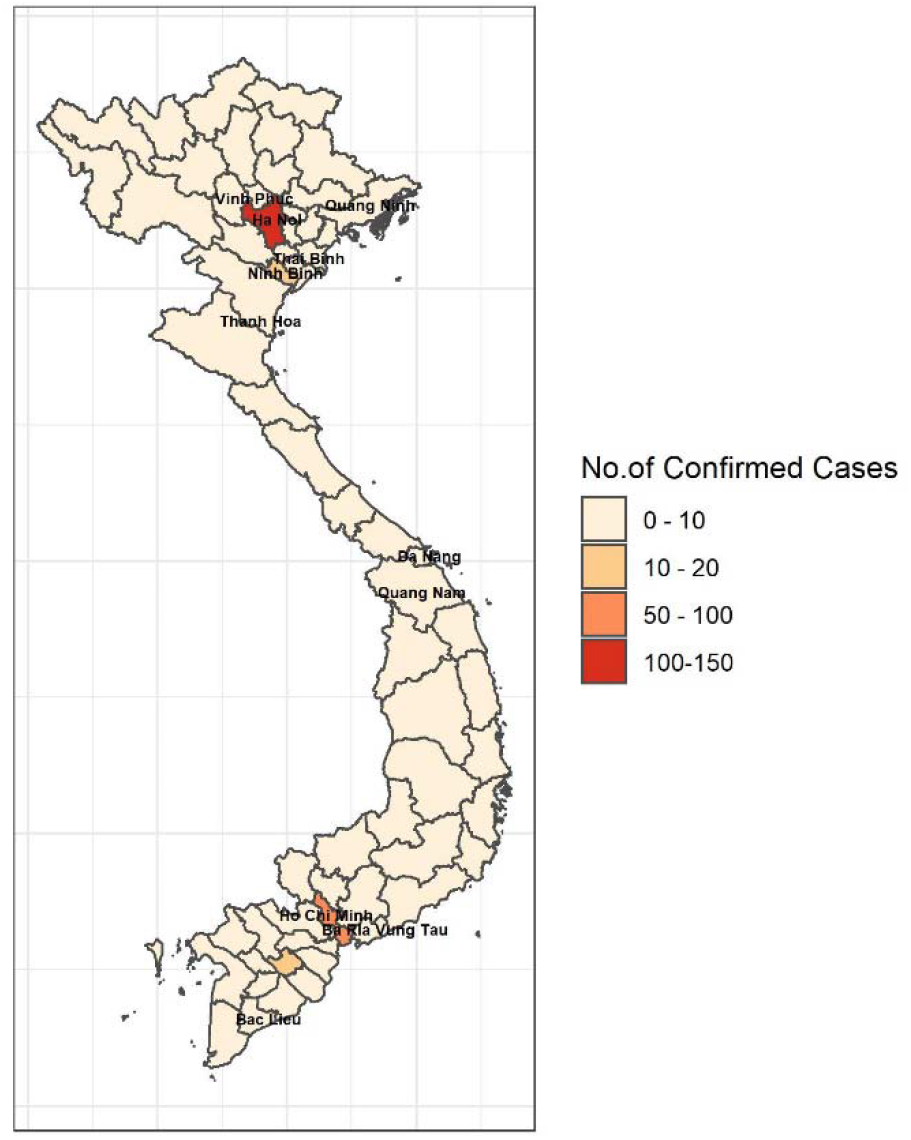
Distribution of Patients with Confirmed Covid-19 Cases across Vietnam, 23 January – 22 April,

**Figure 2c.**
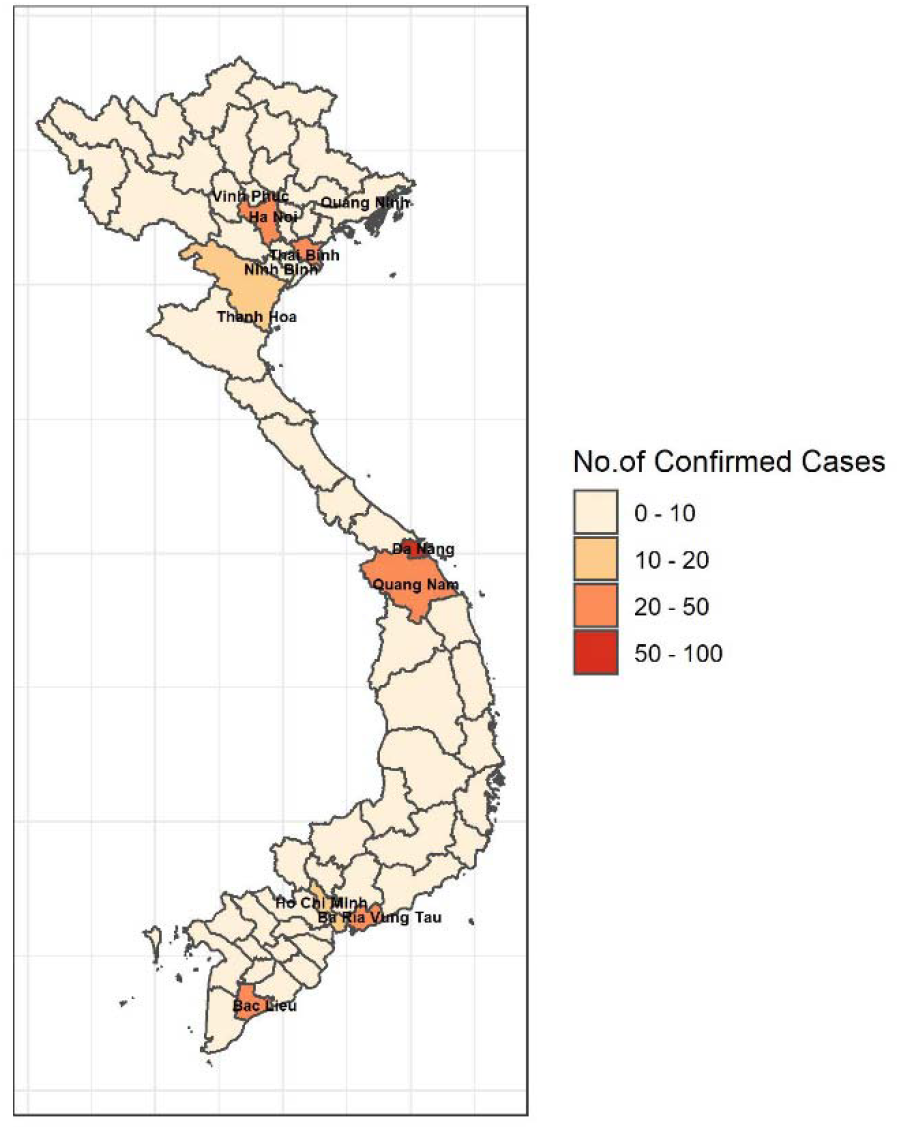
Distribution of Patients with Confirmed Covid-19 Cases across Vietnam, 23 April – 31 July, 2020

The demographic and epidemiological characteristics of the patients are shown in Table 1. Patients had a median age of 38.56 (SD 16.61) years, and the patients in the time after lockdown end were older than those in the period before lockdown end by a median of 5 years. Overall, those aged 21–30 years are recorded with the highest number of Covid-19 confirmed cases in both periods (35.82% (n = 96) vs. 27.91% (n = 84)). Almost 90% (n = 241) of patients aged 59 years or younger before lockdown, while this figure after lockdown end was nearly 85% (n = 254). There was significant difference in the patient gender between two periods, 45.52% male before lockdown and 61.79% male after lockdown.

**Table 1.**
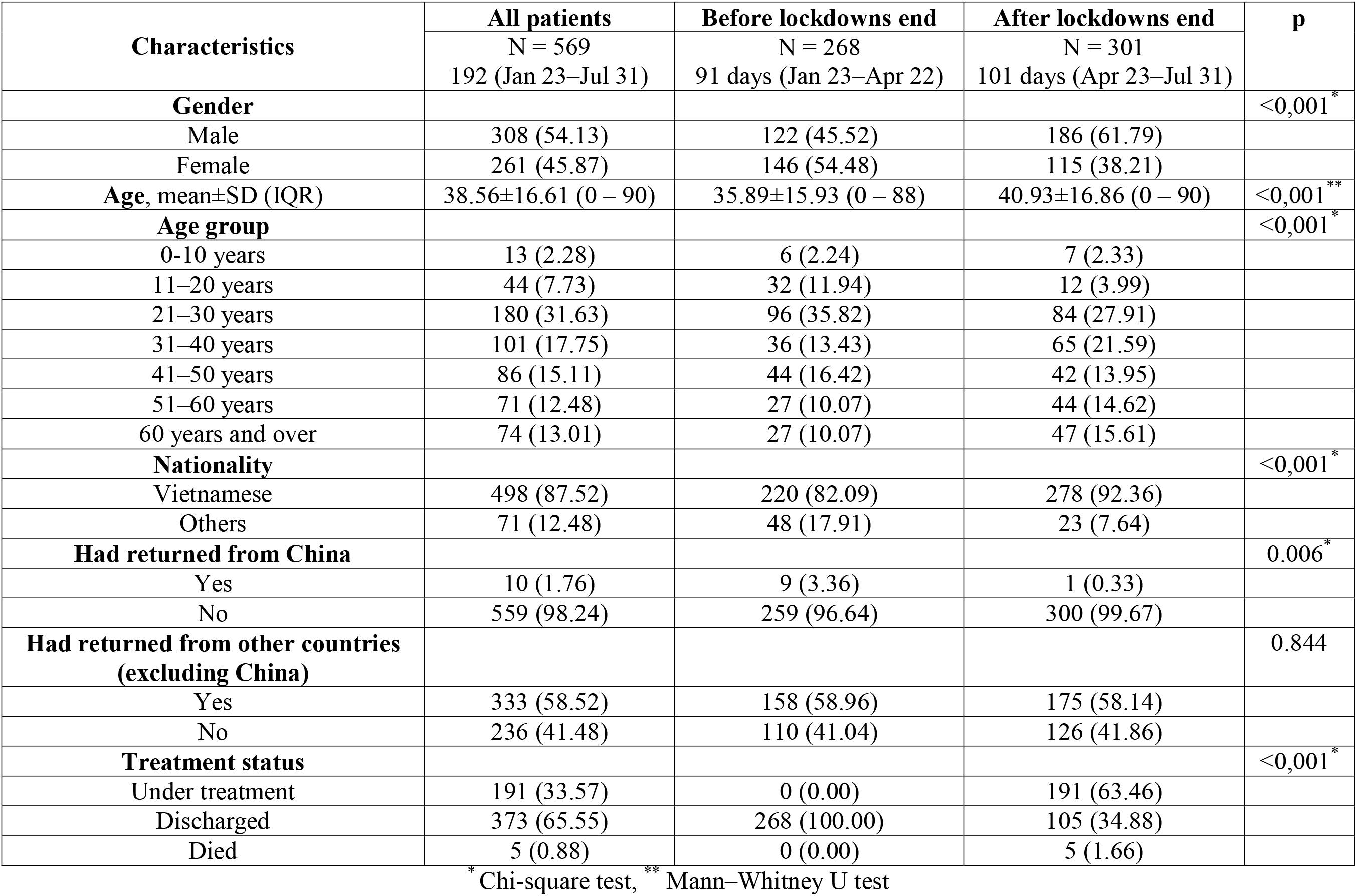
Characteristics of the Covid-19 patients detected before versus after lockdown end, 23 January to 31 July 2020 (N = 569)

Of 10 Covid-19 patients returned from China, 9 patients (3.36%) were in the first period and 1 (0.33%) was in the second period, while no difference between two periods was observed according to the number of patients returned from other countries. Besides, the Covid-19 patient’ nationality between two periods was also significantly varied (Table 1).

Compared to the phase before lockdown with 268 discharged Covid-19 patients (100%) and no Covid-19 death, post lockdown period had only 34.88% patients cured and discharged from the facilities with 5 fatalities (1.66%) and 191 those being under treatment (63.46%) (Table 1).

## Discussion

At the time of writing this article, the first 569 consecutive confirmed cases of Covid-19 through 31 July 2020 are utilized to compare according to two stages, before and after lockdown end in Vietnam. Among the 569 patients, the a median age was 38.56 years, which was significantly lower compared to recent reports in other countries such as 62.2 years in 393 patients in New York City, U.S [14] and 50.1 years in 198 patients in Shanghai, China [15]. Age structure of each country may be the best explanation for the difference in the confirmed case number through two phases. The age distribution of cases was revealed with strong and important role on Covid-19 case fatality [16]. In Vietnam, as compared to before lockdown end, the stage after lockdown had a higher proportion of elder patients. The Covid-19 patient distribution in general is quite similar amongst age groups and those aged 21–30 years are recorded with the highest number of Covid-19 confirmed cases in both periods, implying that SARS-CoV-2 infection risk considerations are not focused solely on any age group. Those in later stage are also patients having pre-existing comorbidities, contributing to the partial explanation of 5 first fatality cases before 31 July. In this analysis, it was observed that significant difference in the gender was shown between two stages (45.52% male patients before lockdown end vs 61.79% male patients after lockdown end).

Despite the more and more increasing global Covid-19 patient number with the overload of the health system, Vietnam has still been responding well to the Covid-19 pandemic with the mobilization of the entire political system. Significant difference was observed in the nationality of Covid-19 patients between two phases. During the initial phase of the study timeline, 17.91% patients (n=48) were residents of other countries, which decreased to 7.64% patients (n=23) during phase after the lockdown end. This point is mainly because, in the time after lockdown end, most of them are Vietnamese citizens flown home from abroad [17]. In our analysis, the Vietnamese cases increase in the community after lockdown may be associated with the reopening of domestic tourism in the “new normal” condition after the first Covid-19 wave and the illegal border entry [4], [18], [19]. Current situation requires that, Vietnam’ proactive efforts in Covid-19 control will be effective when volume, speed, and reach of travel in tourism are well managed and the measures to closely control and monitor repatriation and immigration via its borders are prioritized. Despite having a long borderline with China, Vietnam only recorded 9 confirmed patients returned from China at the time before lockdown end and only a case after lockdown who had illegally immigrated to Vietnam. On the other hand, we found, the number of Covid-19 patients who returned from other countries (excluding China) slightly increased through two stages, however, this difference was not statistically significant. This result partially shows that the continuous volume of people returning Vietnam from abroad during the Covid-19 epidemic. Such the detection of the Covid-19 cases from overseas is the effort of not only the entire political system in immigration control and management of people returning from abroad in general and but also the Vietnam Border Defense Force in particular. The more clear proof is that, over 16,000 people illegally entering Vietnam have been prevented by the Vietnam Border Defence Force since the beginning of 2020 and, particularly, over 2,400 people have been arrested in July once trespassing on the trail [19].

Our study showed that the difference in the treatment status of Covid-19 patients between before and after lockdown end. Vietnam’ early success in the Covid-19 patient treatment was proved by the absolute figure of 268 discharged Covid-19 patients (100%) before lockdown end. Although the number of the recovered patients, overall, continued to slightly increase from 23 April to 25 July after lockdown end, yet the number of new cases in this phase gradually decreased within three months before Covid-19 outbreak point documented in the community in Da Nang on 25 July. Importantly, all new Covid-19 instances from 23 April to 25 July after lockdown were Vietnamese people who returned from abroad and this risk group was then required to isolate for 14 days [20]. When Vietnam was retaining 99 days without any cases in the community, various countries across the world have been faced a rapid growth in the number of new and fatal cases of Covid-19 [21]. Since 25 July after lockdown, almost patients being treatment in Vietnam have been confirmed SARS-CoV-2 infection, which known as the second wave of Covid-19. In such emerging Covid-19 situation, the community transmission cases reported to be linked to the hospitals in Da Nang where the elder patients with chronic severe diseases such as cancer, chronic kidney disease and dialysis, heart failure, diabetes, or hypertension were being treated [22]. Therefore, compared to the phase before lockdown end (all discharged Covid-19 patients (100%) and no death), the post-lockdown time recorded the worse situation with mostly patients being treated (n = 191, 63.46%) and first 5 fatalities (1.66%). It should be noted that five fatal cases were identified at the moment of Vietnam’ second Covid-19 outbreak after the national lockdown implementation for three months. After synthesizing current analysis data, two main reasons associated with Vietnamese Covid-19 deaths were suggested by the Vietnamese experts. Firstly, the new coronavirus strain detected in second wave of Covid-19 infections there had a faster speed of infection compared to the earlier strain. However, many uncertainties have remained concerning the virus-host interaction and the evolution of the epidemic, and its harmfulness compared to the strain previously existed in Vietnam has been not yet known [22], [23]. Secondly, the presence of multiple coexisting severe illness was reported in Covid-19 fatalities [24].

The present results should be viewed with the limitations. First, because the paper’ secondary analysis data was collected from two publicly available datasets, the information of the exposure history, clinical symptoms or signs, radiologic assessments, laboratory testing as well as the other risks could be unknown. Second, the challenges in national large-scale coronavirus testing resulted in the underestimated true number of Covid-19 infections. Finally, due the ongoing Covid-19 pandemic with the more complicated situation in Vietnam, the current estimates are temporary at the time of this writing.

## Conclusions

Our short communication illustrated demographic and epidemiological disparity of Covid-19 patients before versus after loosening the national lockdown in Vietnam. The Covid-19 patient distribution in general is quite similar amongst age groups and these patients were largely recorded in the 21–30-year-old group in both periods. Most of the Covid-19 patients are residents of Vietnam and Vietnamese patient number has still increased rapidly during phase after the lockdown end. Importantly after the lockdown period, Vietnam’ proactive efforts in Covid-19 control after lockdown end will be effective when the measures to closely control and monitor repatriation and immigration via its borders are strictly enforced.

## Data Availability

All data are available on reasonable request.

## Author Contributions

Conceptualization: H-LV; Data analysis: K-DV; Methodology: H-LV and K-DV; Supervision: H-LV; Writing – original draft: H-LV and K-DV; Writing – review and editing: H-LV and KDV.

## Acknowledgments

We thank the Vietnam Ministry of Health for publicly providing the open access Covid-19 databases.

